# Annotation-preserving machine translation of English corpora to validate Dutch clinical concept extraction tools

**DOI:** 10.1101/2024.03.14.24304289

**Authors:** Tom M Seinen, Jan A Kors, Erik M van Mulligen, Peter R Rijnbeek

## Abstract

**Objective:** This work aims to explore the feasibility of validating Dutch concept extraction tools using annotated corpora translated from English, focusing on preserving annotations during translation and addressing the challenge posed by the scarcity of non-English corpora in clinical settings.

**Materials and methods:** Three annotated corpora were standardized and translated from English to Dutch using two machine translation services, Google Translate and OpenAI GPT-4, with annotations preserved through a proposed method of embedding annotations in the text before translation. The performance of two concept extraction tools, MedSpaCy and MedCAT, was assessed across the corpora in both Dutch and English.

**Results:** The translation process effectively generated Dutch annotated corpora, allowing the concept extraction tools to perform similarly in both English and Dutch. Although there were some differences in how annotations were preserved across translations, these did not affect extraction accuracy. Supervised MedCAT models consistently outperformed unsupervised models, whereas MedSpaCy demonstrated high recall but lower precision.

**Discussion:** Our validation of Dutch concept extraction tools on corpora translated from English was successful, highlighting the efficacy of our annotation preservation method and the potential for efficiently creating multilingual corpora. Further improvements and comparisons of annotation preservation techniques and strategies for corpus synthesis could lead to more efficient development of multilingual corpora and more accurate non-English clinical concept extraction tools.

**Conclusion:** This study has demonstrated that translated English corpora can be effectively used to validate non-English concept extraction tools. The annotation preservation method used during translation proved effective, and future research should aim to extend this corpus translation method to additional languages and clinical settings.

## Introduction

Electronic Health Records (EHRs) have become an invaluable source of real-world data for observational research, offering insights into disease prevalence, patient outcomes, and treatment effectiveness [1,2]. While structured data, such as coded conditions, measurements, and prescriptions, are frequently used for analysis, a significant portion of valuable patient information remains locked within free text, such as nursing and physician notes [3,4]. The extraction of information from this unstructured data in a structured manner, in the form of standardized clinical concepts, e.g., from the Unified Medical Language System (UMLS)[5], can greatly enhance observational research by providing additional rich, detailed clinical information at scale [4,6,7]. Numerous tools for this natural language processing (NLP) task of clinical concept extraction, which consists of both named entity recognition and entity linking, have been developed for English clinical texts [8-10], including tools such as cTAKES [11], MetaMap [12], QuickUMLS [13], and MedCAT [14], cloud-based tools [15], and tools using generative large language models [16].

However, the need for concept extraction tools and validating these tools extends beyond English [10], particularly with the rise of real-world data utilization in observational clinical research across the multilingual continent of Europe [17], as seen in initiatives like the European Medical Information Framework (EMIF) [18], the European Health Data & Evidence Network (EHDEN) [19], and the Data Analytics and Real World Interrogation Network (DARWIN EU)[20]. Utilizing unstructured data in large-scale analyses within standardized frameworks, such as the Observational Medical Outcomes Partnership Common Data Model (OMOP CDM), highlights the importance of reliable information extraction for different languages. Nevertheless, the landscape of concept extraction tools for relatively small non-English languages such as Dutch, remains underdeveloped, and the limited number of tools currently available for Dutch clinical text, including adapted versions of QuickUMLS [6] and MedCAT [21], have not been publicly evaluated. At the same time, it is not uncommon for extraction tools to lack validation [22], most English extraction tools are validated using various public corpora annotated with clinical concepts, for example, i2b2 [23], ShARe/CLEF [24], and MedMentions [25]. While benchmarks exist for various other Dutch NLP tasks [26], the absence of Dutch annotated clinical corpora poses a significant challenge for validation and comparison of the extraction tools in this language [27,28].

Creating an annotated clinical corpus in any language is resource-intensive, requiring significant labor to manually annotate numerous clinical texts in great detail [10,29]. The use of pre-trained large language models (LLMs) for data augmentation and generation to create new corpora has been proposed as an alternative to the manual annotation effort [30-32]. For instance, a recent study demonstrated that LLM data generation can produce clinical texts in German, with entities annotated according to broad semantic categories [33]. Besides synthesizing new data, a scalable option that relies on the models’ creativity and domain knowledge, LLMs also enable data augmentation, notably by translating existing English corpora into other languages [32,34]. While machine translation using LLMs has significantly improved in recent years [26,35], merely translating the clinical texts of an annotated corpus is insufficient because the word locations of clinical entities within the text shift during translation, causing the loss of annotation information tied to specific text locations [36]. Although these annotations could be manually repositioned or aligned using secondary word alignment software after translation [34,36,37], we propose a method that preserves annotation locations during translation by embedding the annotations within the text before translation and retrieving them afterward.

Our study investigates the feasibility of validating non-English, specifically Dutch, concept extraction tools using English-annotated corpora translated via machine translation with embedded annotations. We evaluate two English concept extraction tools that were adapted to Dutch, on two English annotated corpora and their Dutch translations, and a multilingual annotated corpus. We compare the concept extraction performance of the tools between the languages.

## Materials and methods

### Experimental setup

The experimental setup consisted of two main parts. The first part involved the corpus translation and preparation phase, where three publicly available annotated corpora were standardized to the same format. This included translating English corpora into Dutch while preserving annotations and creating training and test sets. The second part involved applying and evaluating two concept extraction tools on the test sets, with one tool that supported supervised training, also using the training sets. The setup is visualized in Figure 1.

**Figure 1.**
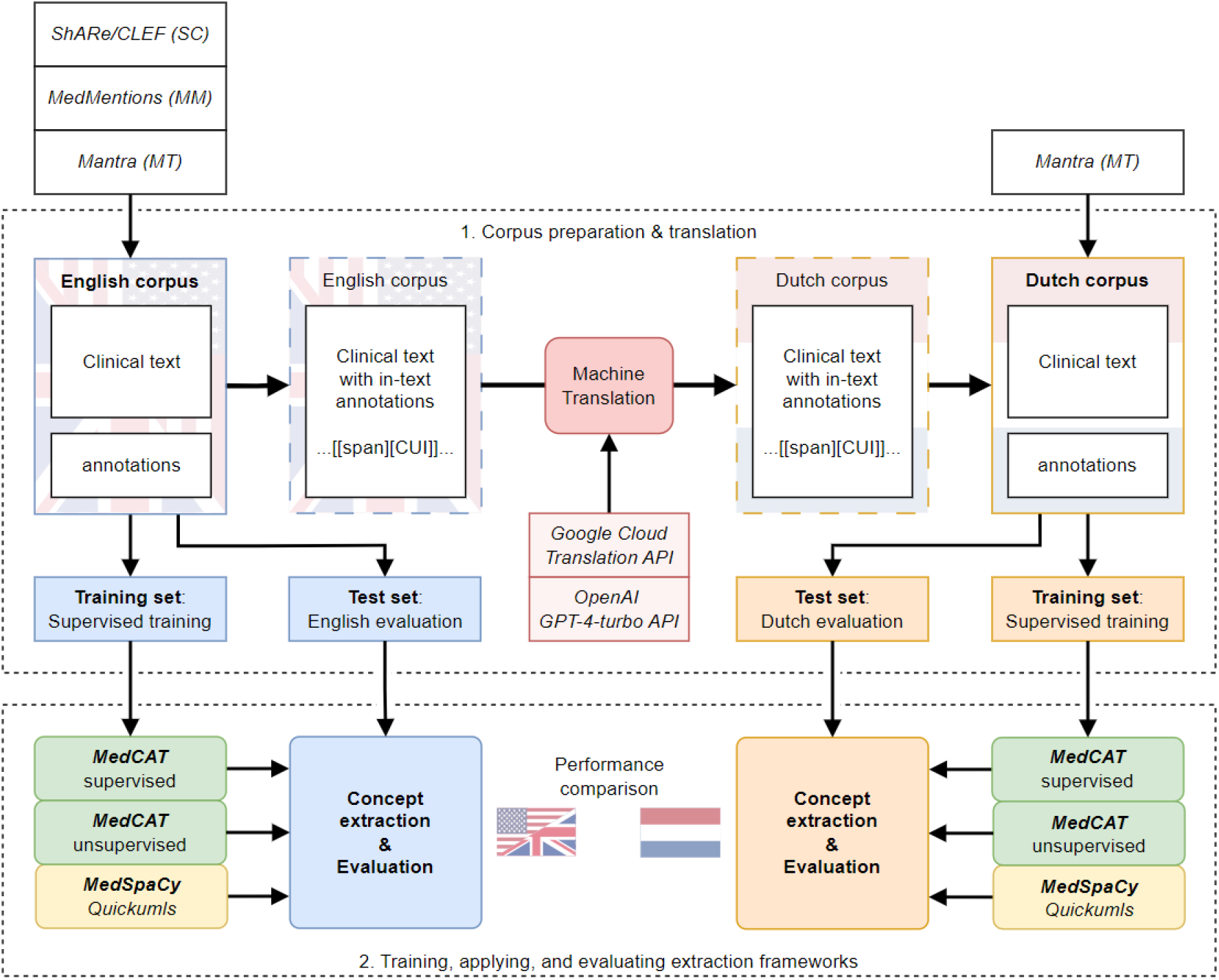
Schema of the experimental setup: 1. preparation and translation of three different corpora (ShARe/CLEF, MedMentions, Mantra), and 2. training, application, and evaluation of two concept extraction tools (MedCAT, MedSpaCy). CUI: concept unique identifier.

### Corpora

The annotated corpora used in our study include the *MedMentions* corpus (MM) [25], the corpus from the *ShARe*/*CLEF eHealth evaluation lab task 2* (SC) [24], and the multilingual *Mantra* corpus (MT) [27]. MM is a comprehensive biomedical corpus containing 4,392 abstracts from PubMed, annotated with concepts across a wide range of biomedical fields. SC is a corpus derived from 432 clinical notes and is designed to facilitate tasks related to understanding clinical text, including entity recognition and normalization. The multilingual MT corpus provides annotations of 200 short texts from different parallel corpora (Medline abstract titles (MDL) and sentences of drug labels from the European Medicines Agency (EMA)), in multiple languages, including English and Dutch. All three corpora feature annotations that link text spans to a UMLS concept, identified by a Concept Unique Identifier (CUI). To facilitate uniform analysis, all corpora were standardized into the same tabular format. This involved separating the text documents (Attributes: DocumentId, Text) and the concept annotations (Attributes: DocumentId, CUI, SpanStart, SpanEnd, SpanText). The SC corpus is pre-partitioned into training and test sets, whereas for MM and MT, we randomly allocated 80% of the data for training and 20% for testing.

### Corpus translation

To develop the Dutch corpora of annotated clinical texts, we used the English annotated corpora as a starting point. Directly translating an English text would allow us to create a Dutch text, but the exact locations of the annotations would be lost. To address this, we propose a method for preserving the location of annotated concepts through a process of three steps (see Table 1 for an example). First, annotations are integrated directly into the clinical text by enclosing the text span and the CUI in square brackets, i.e. [[text span][CUI]]. Next, this text with embedded annotations is translated using machine translation, keeping the annotations intact. Finally, the annotations are extracted from the translated text using a simple regular expression pattern (“\[\[([^\]\[]*)\]\[(C[0-9]*)\]\]”), resulting in separate text documents and annotations once more.

**Table 1.**
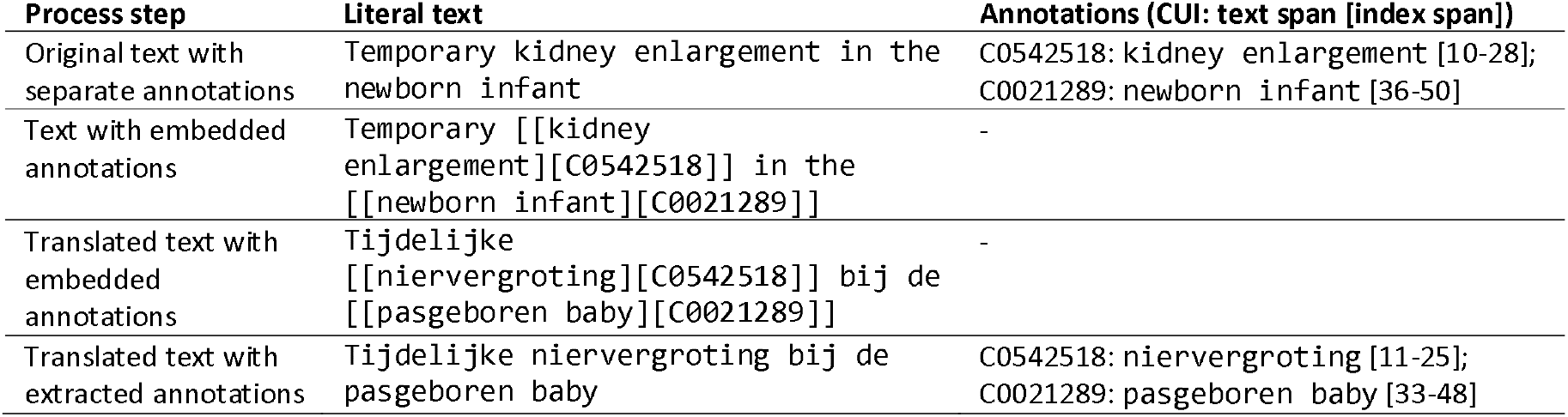
Example phrase from the MT corpus to illustrate the steps in the in-text annotation translation process.

To experimentally assess the impact of translation in this process, we utilized and compared two different machine translation services: the Cloud Translation API from Google (referred to as Google) and the GPT-4 Turbo (gpt-4-0125-preview) API from OpenAI (referred to as GPT) [38]. Google’s service offered direct machine translation of documents. In contrast, GPT, a generative text model, required a specific system prompt besides the document text to guide a zero-shot translation process:

> *“Translate the document to Dutch (Nederlands). Keep the formatting the same, including the in-text annotations: [[span][code]]*.*”*

This approach allowed us to compare a traditional translation service with a state-of-the-art generative text model in preserving annotated concept locations during translation. To evaluate the quality of annotation preservation, we compared descriptive statistics like document size and the number of annotations before and after translation. Additionally, we quantified formatting errors, i.e., the erroneous placing of brackets in the translated text, by counting the CUIs in the final translated text, as an annotation with a wrong bracket pattern is not extracted, and its CUI remains in the text.

### Concept extraction tools

In our study, we validated and compared two concept extraction tools: MedSpaCy^1^ and the Medical Concept Annotation Toolkit^2^ (MedCAT). These Python tools, both open source and publicly available, were initially designed for extracting concepts from English texts and have been adapted for Dutch [6,21].

MedSpaCy extends the spaCy software library for clinical NLP tasks, including clinical concept extraction, using an adaptation of QuickUMLS—a tool for fast, unsupervised biomedical concept extraction based on string similarity and a reference concept dictionary. For English concept extraction, we utilized all UMLS concepts with English terms. For Dutch concept extraction, we used all Dutch vocabularies from UMLS and replaced the English Systematized Nomenclature of Medicine Clinical Terms (SNOMED CT) vocabulary with the Dutch SNOMED CT translation^3^, maintained by NICTIZ, the Dutch National IT Institute for Healthcare^4^. If no Dutch version of a UMLS vocabulary existed, we kept the English one, as many concepts, such as drug and laboratory concepts, are language-independent. Further details on QuickUMLS settings are provided in Supplementary Table S1, and detailed information on the concept dictionaries is available in Supplementary Table S2.

MedCAT is an entity recognition and linking tool, employing context similarity based on word embeddings for concept recognition and disambiguation [14]. It allows for both unsupervised training on unannotated clinical texts and supervised training on annotated texts. In this study, we used the publicly available pre-trained models for English and Dutch, developed using unsupervised training. The English model, trained on clinical notes from Medical Information Mart for Intensive Care (MIMIC) III [39], used a subset of the English UMLS as its concept dictionary. The Dutch model is trained on medical Wikipedia articles in Dutch, incorporating all UMLS concepts with Dutch descriptions and the Dutch SNOMED CT translation [21]. Additionally, to showcase MedCAT’s supervised training capabilities, we fine-tuned the pre-trained unsupervised models with supervised learning, creating a separate supervised MedCAT model on the training set of each corpus.

To summarize, our analysis involves three types of concept extraction models: one MedSpaCy model and one unsupervised MedCAT model for both languages, plus ten supervised MedCAT models—three for English for each corpus and seven for Dutch, corresponding to each corpus translation.

### Evaluation

All models were applied to their respective test sets in both English and Dutch. We evaluated the extraction performance using precision, recall, and their harmonic mean, the F1 score [25]. A concept was considered correctly extracted if both the CUI and its location were accurately identified, all other predicted concepts were counted as false positives, and all unmatched reference concepts were counted as false negatives. Furthermore, to compare the overall performance across the languages and the concept extraction models, we mean-centered the evaluation results by the individual corpus. This involved subtracting the corpus mean metric value from each result, allowing for a comparison of extraction outcomes that is independent of the specific corpora. The statistical significance of the differences in metric value distribution was assessed using Bonferroni-adjusted Wilcoxon tests.

## Results

### Corpus translation

The three corpora, MM, SC, and MT, were processed and translated using the two machine translation services, Google and GPT, while preserving the annotations. Characteristics of the corpora and their translations are presented in Table 2. MM was the largest corpus with also the highest annotation density, containing many annotations in a relatively short text, especially compared to the SC corpus. The translation of English to Dutch (with the annotations extracted) increases the number of characters, which is also visible in the existing Dutch translation in the multilingual MT corpus.

**Table 2.** Characteristics of the original English and the Dutch translated corpora.

| Characteristics | Language | MM | SC train | SC test | MT EMA | MT MDL |
| --- | --- | --- | --- | --- | --- | --- |
| No. of documents | - | 4392 | 299 | 133 | 100 | 100 |
| No. of unique UMLS semantic types | - | 126 | 11 | 11 | 73 | 73 |
| Median no. of characters per document | English | 1493 | 2610 | 7370 | 97.5 | 54.5 |
|  | Google | 1742 | 2764 | 7840 | 111 | 58.5 |
|  | GPT | 1769 | 2727 | 7850 | 108.5 | 57.5 |
|  | Dutch | - | - | - | 115 | 59.5 |
| Median no. of annotations per document | English | 79 | 31 | 54 | 3 | 2 |
|  | Google | 79 | 31 | 54 | 3 | 2 |
|  | GPT | 75 | 31 | 52 | 3 | 2 |
|  | Dutch | - | - | - | 3 | 2 |
| Total no. of annotations | English | 352496 | 8381 | 11937 | 363 | 222 |
|  | Dutch | - | - | - | 364 | 214 |
| Total no. of missing annotations<br>(% of annotations) | Google | 2877 | 54 | 52 | 6 | 0 |
|  |  | (0.8%) | (0.6%) | (0.4%) | (1.7%) | (0.0%) |
|  | GPT | 20806 | 232 | 203 | 2 | 0 |
|  |  | (5.9%) | (2.8%) | (1.7%) | (0.6%) | (0.0%) |
| No. of annotations missing due to formatting errors<br>(% of missing annotations) | Google | 2753 | 51 | 29 | 6 | 0 |
|  |  | (95.7%) | (94.4%) | (55.8%) | (100.0%) | (0.0%) |
|  | GPT | 123 | 12 | 4 | 1 | 0 |
|  |  | (0.6%) | (5.2%) | (2.0%) | (50.0%) | (0.0%) |

The quality of in-text annotation preservation was measured by the number of missing annotations after the translation and how many of these annotations were missing due to formatting errors during translation. Overall, the preservation of annotations during translation was quite effective: the median number of annotations per document was the same or close to the original English version. However, GPT translations exhibited the highest percentage of missing annotations, with 5.9% in MM and 2.8% in SC. Google translations performed better, with less than 1% missing annotations, most of which could be attributed to formatting errors. In contrast, GPT showed a very low rate of formatting errors, and its missing annotations were primarily due to the pure loss of embedded annotations during translation: annotations were ignored in the generated text while keeping the sentence structure intact. Table 3 presents examples of both types of annotation preservation errors. Upon further inspection of the missing annotations in the GPT translation, we found that GPT primarily struggled with annotations related to verbal phrases or generic nouns, such as “investigates,” “performed,” “comparison,” and “evaluation.” These missing concepts were predominantly categorized under the more generic semantic types like Functional Concept (3715 concepts, 18%), Qualitative Concept (3593, 17%), and Activity (1591, 8%), while semantic types related to diseases and medicine were barely affected. For a detailed breakdown of the missing annotations in GPT’s MM translations by semantic type and its most frequent unpreserved concepts, see Supplementary Table S3.

**Table 3.**
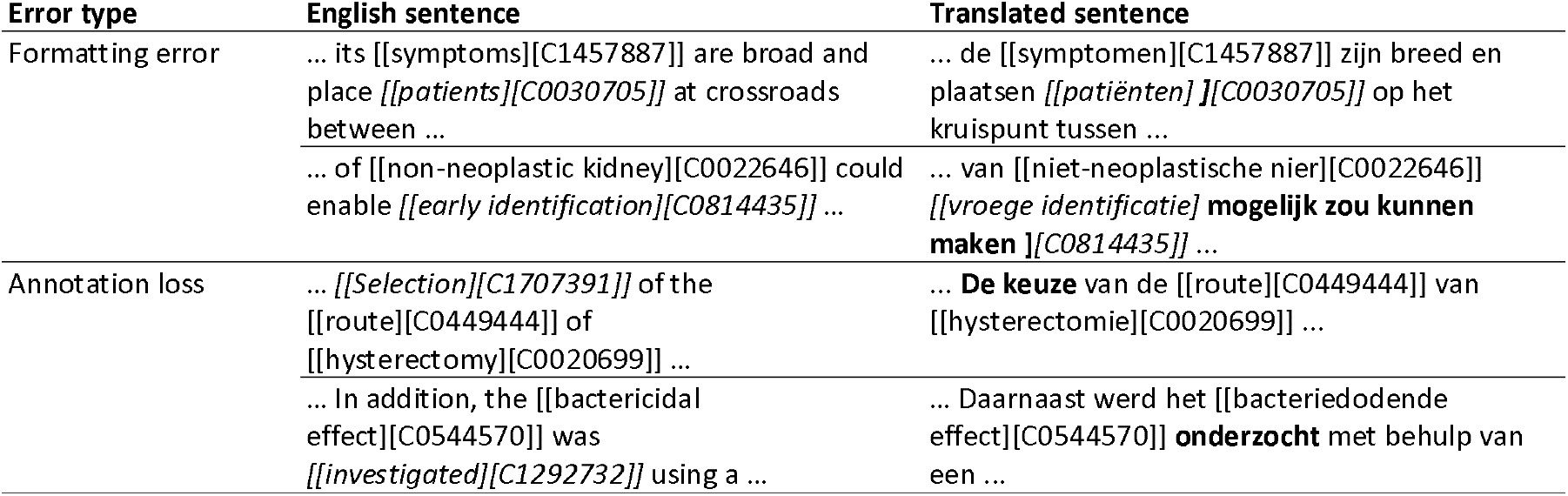
Examples of sentences from the MM corpus with formatting errors and the loss of annotations. The text in italics indicates the erroneous annotation, and the bold text indicates the problem. The sentence translations are correct.

### Concept extraction

In total, 30 model and corpus combinations were evaluated. The performances, measured by the F1 score (F), recall (R), and precision (P), are visualized in Figure 2. For the exact values, see Supplementary Table S4. Overall, the concept extraction tools performed similarly in English and translated Dutch corpora. Despite several differences between English and Dutch within each corpus, we observed, on average, that there were no significant language differences across the different corpora (Figure 3A). Additionally, there were no performance differences between models using Dutch Google or GPT translations or between MT translations and the existing Dutch MT corpus.

**Figure 2.**
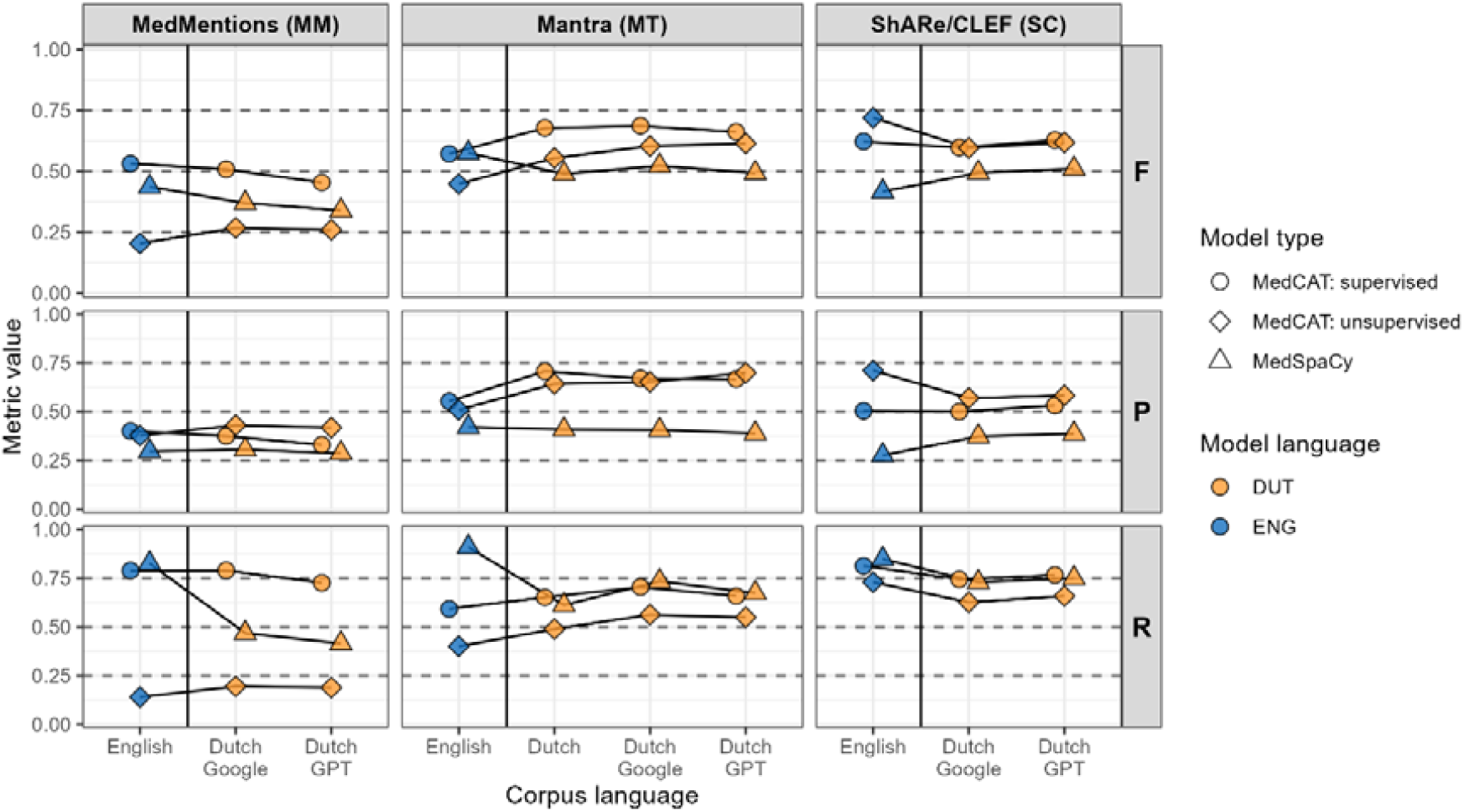
Concept extraction performance per model type and corpus combination on the English version (blue) and on the (translated) Dutch versions (orange) of the three main corpora, measured by the three metrics: F1 score (F), precision (P), and recall (R).

**Figure 3.**
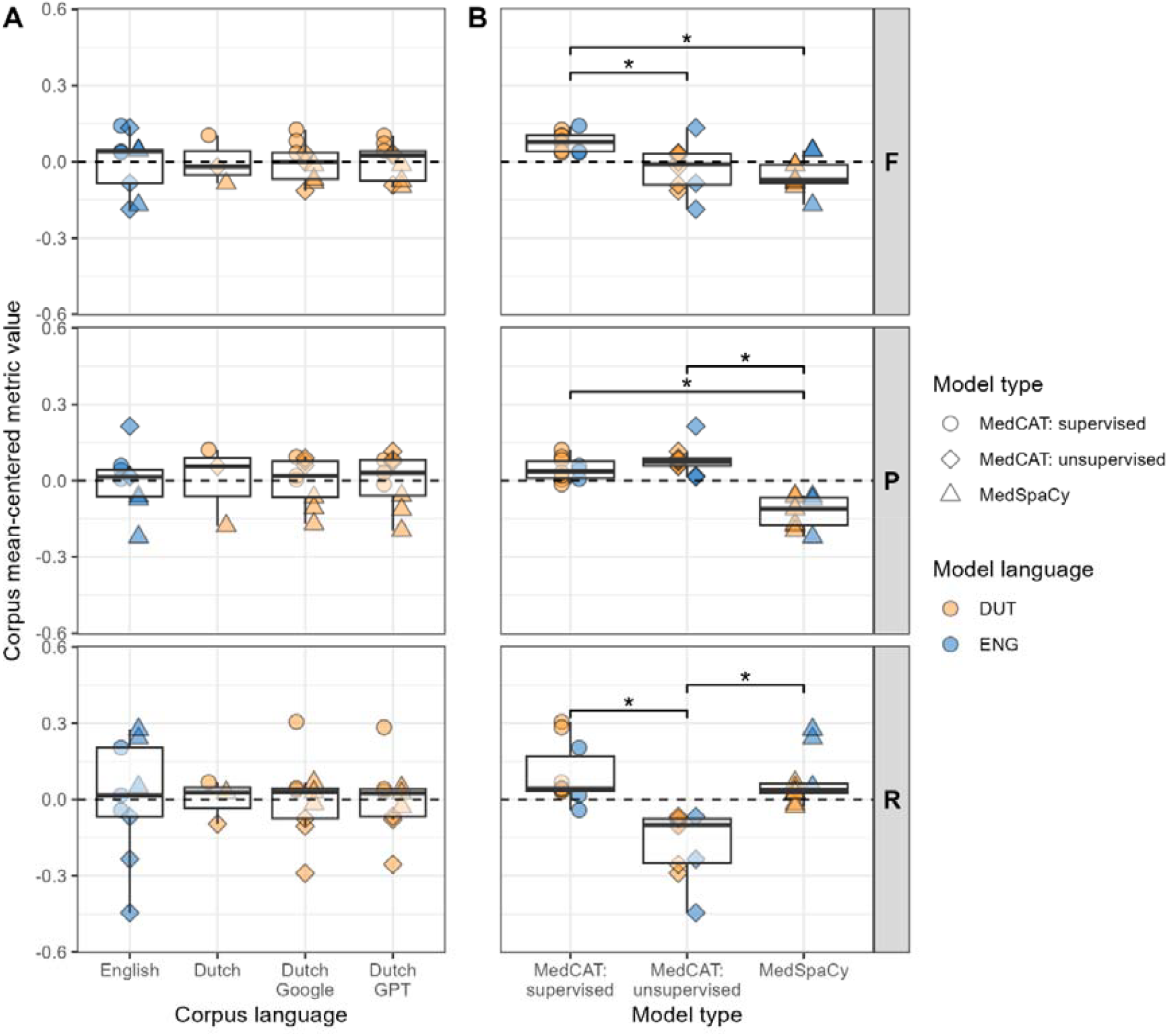
Performance comparison of (A) the different corpus languages and (B) the different concept extraction models independent of the corpora, using mean-centered metric value distributions. The dashed line indicates the mean center. The significant Bonferroni-adjusted Wilcoxon test results between the distributions are shown above the boxplots if significant, where “*” indicates a P value <0.01. The points represent the underlying data.

Within each corpus, performance differences could be observed between the concept extraction model types. In the MM corpus, the supervised MedCAT model performed the best, followed by MedSpaCy, in both languages. The differences in performance could mainly be attributed to the large differences in recall. Notably, the low recall of the unsupervised MedCAT models showed a drastic improvement with supervised learning. Furthermore, the high recall of the English MedSpaCy was not mirrored in the Dutch version, possibly due to the wide semantic type range in MM and the lower number of concepts with Dutch terms in the Dutch MedSpaCy. In the MT corpus, we also found that the supervised MedCAT model had the best performance, with a similar performance of MedSpaCy in English, followed by unsupervised MedCAT in Dutch. Again, the high performance of MedSpaCy in English was attributed to its high recall, as precision differences were relatively small. Conversely, in the SC corpus, the differences between the model types were mainly due to differences in their precision. Both unsupervised and supervised MedCAT models performed similarly, with the unsupervised model slightly outperforming the supervised one in the English corpus. Although supervised training improved recall, it reduced precision. Figure 3B presents the mean-centered performance across corpora, showing that, on average, supervised models performed best. While MedSpaCy models had a high recall similar to supervised MedCAT, their precision was consistently lower.

## Discussion

### Concept extraction evaluation using translated corpora

This study explored the feasibility of validating Dutch concept extraction tools on annotated corpora derived from translating existing English corpora. We validated two concept extraction tools in Dutch and English using one annotated multilingual corpus and two annotated English corpora. The results demonstrated the effective generation of Dutch annotated corpora through our proposed method, which preserves annotation location through translation, facilitating rapid, efficient, and accurate creation of Dutch corpora annotated with clinical concepts, without necessitating further post-processing for text alignment [34].

We successfully utilized machine translation services, Google and GPT, for corpus translation. Google encountered more issues with annotation formatting, whereas GPT translations had a larger number of missing annotations, primarily due to problems with verbal phrases and generic nouns, affecting the preservation of annotations. This issue was particularly notable in the MM corpus, which has a high annotation density. The exact reason for these missing annotations is unclear but may be related to these phrases and nouns not typically annotated as clinical entities.

The translation process from English to Dutch did not significantly impact the performance of concept extraction, with models showing comparable effectiveness across languages and corpora. Moreover, no significant differences were observed in model performance between Google or GPT-translated corpora or between the Dutch MT corpus and the MT translations. These results confirm the feasibility of accurately translating existing annotated corpora for multilingual use, as demonstrated by the method’s effectiveness for Dutch, which is broadly applicable and expected to perform well across various languages.

When comparing the performance of the concept extraction models across the different corpora, we found that the supervised MedCAT model generally performed best. The fine-tuning of the unsupervised models using supervised learning showed much improvement, especially in the MM and MT corpora. These findings for MedCAT align with those reported by the authors [14]. MedSpaCy demonstrated a high recall across all corpora but suffered from lower precision, likely due to its reliance on an extensive concept database that led to the extraction of many correct concepts alongside numerous unannotated ones.

Overall, this research enhanced our understanding of the challenges and opportunities in creating multilingual annotated clinical corpora and validating non-English concept extraction tools, contributing to clinical NLP and data harmonization to improve observational research.

### Strength and limitations

Although the three corpora used in this study span different settings, including clinical notes from an American hospital EHR, biomedical scientific abstracts, and European drug labels, in practice, Dutch clinical text might differ from its English language counterpart due to variances in healthcare systems and practices. Thus, while translating existing English corpora provides a rapid method for generating new corpora in other languages, creating new corpora in the target language and, crucially, within the target setting remains preferable.

For corpus translation, we relied on two leading LLM services used for translation without systematically validating the accuracy of the resulting translations. However, we identified no significant issues through ongoing observation and comparison during the study. Despite this, machine translation is not infallible, and nuances may be lost, potentially impacting the reliability of the annotated corpus. To address this, we assessed and quantified errors in the annotation-preserving translation process using simple metrics and published details on missing annotations for public scrutiny. Our choice of the Google Cloud Translation API and the OpenAI GPT-4 API allowed us to compare a dedicated machine translation model with a large generative model not explicitly designed for translation. With GPT, we only employed zero-shot prompting and observed good results despite occasional annotation losses and minimal formatting errors. However, GPT allows further improvement through techniques like prompt optimization and few-shot learning, highlighting its versatility. The Google translations exhibited more annotation formatting errors than GPT. While the Google translation model cannot be directly altered, addressing the errors with more complex regular expressions would further improve the annotation preservation.

We validated and compared a limited number of concept extraction tools chosen for their ease of use and availability in Dutch and English. Although we used default settings for the MedSpaCy and MedCAT models, further optimization could enhance performance. The performance of the concept extraction models was not optimal, with F1 scores of the best models ranging between 0.5 and 0.7 across corpora. However, our stringent evaluation required an exact match of the predicted and annotated CUI to be considered correct [25]. We observed that many predicted concepts closely matched the annotated concepts and, in some instances, could be considered more accurate. For instance, the word “seizure” is annotated as C0036572: Seizures, but the model extracts the similar C4229252: Seizure-like activity. Similarly, the phrase “cocaine use” is annotated as C0009171: Cocaine Abuse, but the model extracts C3496069: Cocaine Use. Therefore, a less strict evaluation method based on close concept similarity, measured by hierarchical or concept embedding distance, would likely yield higher performance.

### Future work

Future work should explore the generalization of our corpus translation method to languages beyond Dutch. While translating existing corpora offers an efficient alternative to creating new ones from scratch, comparing this method to others, such as exploring synthesizing corpora using LLMs to generate new CUI annotated data based on examples or combining multiple strategies, would be worthwhile. Our annotation preservation technique shows promise, but further research is needed to optimize its accuracy. Improvement could involve experimenting with various LLM models, employing one-shot or few-shot prompting, using more extensive prompts with more instructions, and fine-tuning models. Additionally, different methods for embedding annotations in the text, such as using curly or angle brackets or even complete JSON, HTML, or XML markups, could be explored. A comparative analysis of our annotation preservation method with post-translation word alignment techniques, as proposed by others [34], would also be valuable. Lastly, others can use the translated corpora from our study to evaluate different concept extraction tools, train models, and adapt our translation approach for translating other corpora in various clinical settings and NLP tasks.

## Conclusion

In conclusion, this study demonstrated the feasibility of validating Dutch concept extraction tools using annotated corpora translated from English. The proposed method of preserving in-text annotations during translation through language models offers a promising alternative to post-translation realignment of words. The research extended to three different corpora, two machine translation services, and two extraction tools, showcasing the method’s versatility and potential for multilingual clinical NLP advancement. While machine translation services like Google and GPT were effective in translating annotated clinical corpora, some issues were encountered, highlighting the need for ongoing optimization and error assessment. The comparison of concept extraction models showed that the supervised MedCAT model generally performed best, with MedSpaCy demonstrating high recall but lower precision. Future work should focus on generalizing the corpus translation method to other languages, optimizing annotation preservation techniques, and exploring different strategies for embedding annotations in text. Comparative analysis with post-translation word alignment techniques and further experimentation with various language models and prompting techniques could also enhance the accuracy and efficiency of concept extraction in multilingual settings. This study contributes valuable insights into expanding clinical data augmentation and concept extraction research into non-English languages, paving the way for more extensive multilingual clinical NLP applications and advancements in the field.

## Data availability

The translated MedMentions and Mantra corpora are made publicly available to enhance transparency and reproducibility and facilitate further research: https://github.com/mi-erasmusmc/DutchClinicalCorpora.

Due to access restrictions and a data use agreement with PhysioNet, the translated ShARe/CLEF corpus cannot be made publicly available. The aggregated data used for generating the results, conclusions, and figures/tables in this study are available as supplementary data.

## Author Statement

T.S. proposed the methodology, designed, and implemented the study protocol, and performed the data analysis. J.K., E.M., and P.R. provided critical feedback, helped interpret the results, and shaped the research and analysis. T.S. wrote the article with valuable input from all other authors.

## Acknowledgments

None to declare.

## Supplementary Material

Supplementary Tables

**Supplementary Table S1.** QuickUMLS settings in each MedSpaCy model.

| Setting | Value |
| --- | --- |
| overlapping_criteria | score (default) |
| threshold | 0.7 (default) |
| window | 10 |
| min_match_length | 3 (default) |
| similarity_name | Jaccard (default) |
| accepted_semtypes | List of UMLS semantic types that was used to annotate each corpus |

**Supplementary Table S2.** Details on the concept dictionaries for the Dutch and English MedSpaCy and unsupervised MedCAT models.

| Statistic | MedSpaCy<br>English | MedSpaCy<br>Dutch | MedCAT: unsupervised<br>English | MedCAT:<br>Unsupervised<br>Dutch |
| --- | --- | --- | --- | --- |
| Description | UMLS<br>version<br>2022AB | UMLS<br>version<br>2022AB | *UMLS Small (A modelpack*<br>*containing a subset of*<br>*UMLS (disorders,*<br>*symptoms, medications...)*<br>*Trained on MIMIC-III)* | *UMLS Dutch v1.10 (a*<br>*modelpack provided by UMC*<br>*Utrecht containing UMLS*<br>*entities with Dutch names*<br>*trained on Dutch medical*<br>*Wikipedia articles and a*<br>*negation detection model*<br>*repository/paper trained on*<br>*EMC Dutch Clinical Corpus).* |
| Total no. of concepts | 4,612,422 | 4,074,856 | 573,284 | 383,628 |
| Total no. of terms | 9,803,663 | 8,354,007 | 1,950,576 | 1,123,689 |
| Total no. semantic types | 127 | 127 | 127 | 114 |

**Supplementary Table S3**. Number of missing annotations in GPT’s MM translations per semantic type, with the top-5 most frequently missed concepts.

CSV file: *TableS3_MM_GPT_missing_concepts_top5_per_semtype.csv*

**Supplementary Table S4.** Concept extraction performance per model type and corpus combination on the English version and on the (translated) Dutch versions of the three main corpora, measured by the three metrics: F1 score (F), precision (P), and recall (R).

| Corpus language | Model type | MedMentions (MM) |  |  | Mantra (MT) |  |  | ShARe/CLEF (SC) |  |  |
| --- | --- | --- | --- | --- | --- | --- | --- | --- | --- | --- |
|  |  | P | R | F | P | R | F | P | R | F |
| English | MedSpaCy | 0.40 | 0.79 | 0.53 | 0.55 | 0.59 | 0.57 | 0.50 | 0.81 | 0.62 |
|  | MedCAT unsupervised | 0.38 | 0.14 | 0.20 | 0.51 | 0.40 | 0.45 | 0.71 | 0.73 | 0.72 |
|  | MedCAT supervised | 0.30 | 0.83 | 0.44 | 0.42 | 0.91 | 0.58 | 0.28 | 0.85 | 0.42 |
| Dutch | MedSpaCy |  |  |  | 0.71 | 0.65 | 0.68 |  |  |  |
|  | MedCAT unsupervised |  |  |  | 0.64 | 0.49 | 0.55 |  |  |  |
|  | MedCAT supervised |  |  |  | 0.41 | 0.61 | 0.49 |  |  |  |
| Dutch (Google) | MedSpaCy | 0.38 | 0.79 | 0.51 | 0.67 | 0.70 | 0.69 | 0.50 | 0.74 | 0.60 |
|  | MedCAT unsupervised | 0.43 | 0.19 | 0.27 | 0.65 | 0.56 | 0.60 | 0.57 | 0.63 | 0.60 |
|  | MedCAT supervised | 0.31 | 0.47 | 0.37 | 0.41 | 0.73 | 0.52 | 0.37 | 0.73 | 0.49 |
| Dutch (GPT) | MedSpaCy | 0.33 | 0.73 | 0.45 | 0.67 | 0.66 | 0.66 | 0.53 | 0.77 | 0.63 |
|  | MedCAT unsupervised | 0.42 | 0.19 | 0.26 | 0.70 | 0.55 | 0.62 | 0.58 | 0.66 | 0.62 |
|  | MedCAT supervised | 0.29 | 0.41 | 0.34 | 0.39 | 0.67 | 0.49 | 0.39 | 0.75 | 0.51 |

## Footnotes

1 https://github.com/medspacy/medspacy

2 https://github.com/CogStack/MedCAT

3 https://github.com/mi-erasmusmc/medspacy_dutch

4 https://www.snomed.org/member/netherlands

